# A bootstrap particle filter for viral *R*_*t*_ inference and forecasting using wastewater data

**DOI:** 10.64898/2026.03.06.26347747

**Authors:** Wenfei Fiona Xiao, Yuke Wang, Nikunj Goel, Marlene Wolfe, Katia Koelle

## Abstract

Wastewater is increasingly being recognized as an important data stream that can contribute to infectious disease surveillance and forecasting. With this recognition, a growing number of statistical inference approaches are being developed to use wastewater data to provide quantitative insights into epidemiological dynamics. However, few existing approaches have allowed for systematic integration of data streams for inference, for example by combining case incidence data and/or serological data with wastewater data. Furthermore, only a subset of existing approaches have been able to handle missing data without imputation and to handle datasets with different sampling times or intervals. Here, we develop a statistically rigorous, yet lightweight, approach to infer and forecast time-varying effective reproduction numbers (*R*_*t*_ values) using longitudinal wastewater virus concentrations either alone or jointly with additional data streams including case incidence data and serological data. Our approach relies on a state-space modeling approach for inference and forecasting, within the context of a simple bootstrap particle filter. We first describe the structure of our underlying disease transmission process model as well as our observation models. Using a mock dataset, we then show that *R*_*t*_ can be accurately estimated by interfacing this model with case incidence data, wastewater data, or a combination of these two data streams using the bootstrap particle filter. Of note, we show that these data streams alone do not allow for reconstruction of underlying infection dynamics due to structural parameter unidentifiability. We then apply our particle filter to a previously analyzed SARS-CoV-2 dataset from Zurich that includes case data and wastewater data. Our analyses of these real-world datasets indicate that incorporation of process noise (in the form of environmental stochasticity) into the state space model greatly improves our ability to reconstruct the latent variables of the model. We further show that underlying infection dynamics can be made identifiable through the incorporation of serological data and that the bootstrap particle filter can be used to make forecasts of *R*_*t*_, case incidence, and wastewater virus concentrations. We hope that the inference approach presented here will lead to greater reliance on wastewater data for disease surveillance and forecasting that will aid public health practitioners in responding to infectious disease threats.

## 1 Introduction

Wastewater-based epidemiology focuses on the analysis of wastewater samples to monitor drugs and/or pathogens (Singer et al., 2023). Pathogen surveillance through wastewater sampling and analysis was initially implemented in the 1930s to monitor polio virus, providing critical insights into where the virus circulated. More recently, the COVID-19 pandemic reinvigorated widespread interest in the use of wastewater-based epidemiology as a valuable tool for tracking infectious diseases (Diamond et al., 2022), particularly of viral pathogens that replicate in the gut. Wastewater-based surveillance of viral pathogens is currently being used in several different capacities. These include: (1) to reveal the circulation of specific pathogens within a community that are rarely detected through case surveillance; (2) to identify viral strains or variants of concern that may be circulating using targeted sequencing of wastewater samples (Karthikeyan et al., 2022); (3) to provide information on patterns of disease circulation within a community (Adams et al., 2024). Monitoring of wastewater thus has the ability to improve viral surveillance and therewith the potential to guide public health responses.

One challenge for relying on wastewater samples for quantifying patterns of disease circulation within a community is a computational one: how can virus concentration measurements be used effectively for infectious disease inference? To address this challenge, a growing number of statistical methods have been developed in the last five years to enable wastewater virus concentration data to be interfaced with infectious disease models for inference purposes (e.g., Huisman et al. (2022); Goldstein et al. (2023); Watson et al. (2024); Champredon et al. (2024)). Recently, analyses have also begun to systematically evaluate and compare the performance of existing inference approaches. For example, a recent study has compared wastewater inference methods through their application to wastewater datasets from New York State treatment plants (Hill et al., 2025). Existing methods generally aim to infer the time-varying effective reproduction number (*R*_*t*_) from wastewater virus concentration data, defined as the average number of secondary infections generated by an infected individual. *R*_*t*_ is time-varying because it is influenced by factors such as changes in population susceptibility and changes in contact rates. Reconstructing patterns in *R*_*t*_ is essential because it allows us to detect changes in disease transmission over time (Gostic et al., 2020) and can thereby provide insight into the effects of both non-pharmaceutical interventions and pharmaceutical measures (e.g., vaccination) on transmission dynamics.

Many of the existing statistical methods for *R*_*t*_ estimation using wastewater data have limitations that can restrict their applicability. These limitations include the need to impute missing data, the ability to interface with only wastewater data (rather than perform joint inference using wastewater data and additional data sets), and the extent to which some of these methods are computationally intensive. Here, we develop a new statistical inference approach for using wastewater data to infer *R*_*t*_ that addresses these limitations. Unlike a previous approach that used wastewater data to first predict infections and then use these predictions to infer *R*_*t*_ (Huisman et al., 2022), our approach instead uses a state-space modeling framework to directly infer *R*_*t*_. Our approach, at its core, combines a semi-mechanistic epidemiological process model with observation models to infer *R*_*t*_ using a bootstrap particle filter. In the development and testing of our approach, we first apply our approach to a simulated (mock) dataset, where we know the true underlying parameters. We then apply our approach to a previously published dataset from Zurich, Switzerland (Huisman et al., 2022) to illustrate its use on real-world data. Finally, in the context of the Zurich dataset, we show how serological data can be used to identify previously unidentifiable parameters and infection dynamics and how this approach can be used in forecasting.

## 2 Methods

### 2.1 The state-space model

State-space models combine process models with observation models for statistical inference (Kantas et al., 2015; Aoki, 1990). We describe our semi-mechanistic process model in Section 2.1.1 and the observation models we use for case incidence data and wastewater data in Section 2.1.2. Table 1 summarizes the variables and parameters used in these models. Our use of a semi-mechanistic process model enables us to incorporate demographic constraints that restrict the flow of information in a biologically meaningful manner.

**Table 1.**
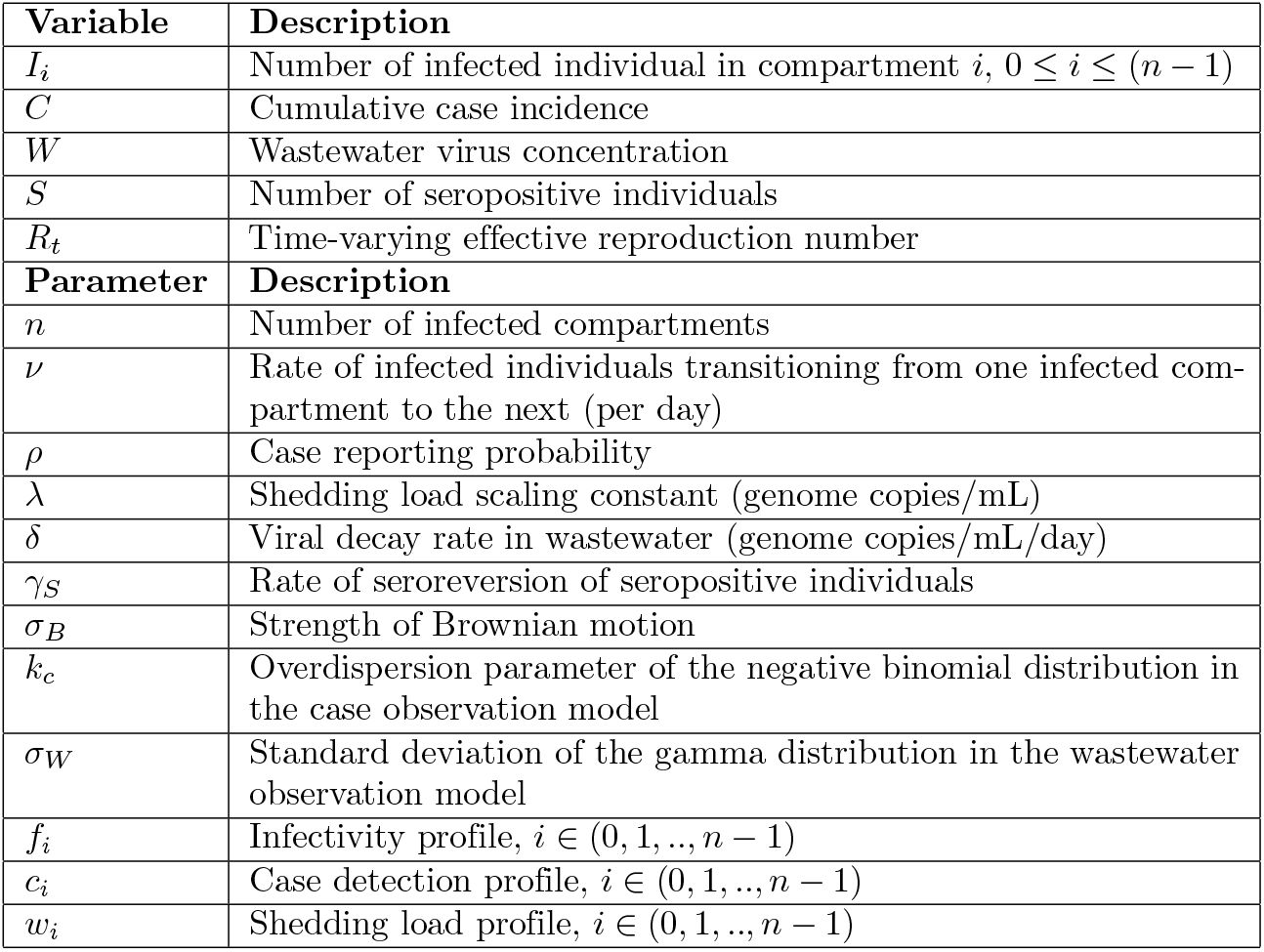
Variables and parameters used in the state-space model.

#### 2.1.1 The process model

Our process model simulates the dynamics of infected individuals, cumulative case incidence, and virus concentrations in wastewater. We partition infected individuals into *n* compartments, with individuals who become infected starting in compartment *I*_0_ and transitioning into downstream compartments over time. The dynamics of infected individuals are given by:

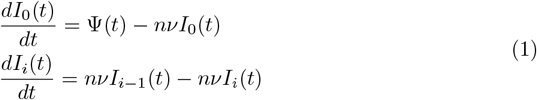

where Ψ(*t*) is a time-varying infection incidence rate and *i* indexes downstream compartments, where 1 ≤ *i* ≤ (*n*− 1). Infected individuals transition from one compartment to the next at a rate of *nν*, such that the time between when an individual becomes infected and when they leave the last infectious class is on average 1*/ν*. The time-varying infection incidence rate is given by:

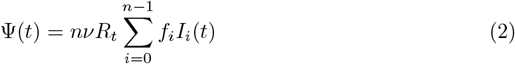

where *R*_*t*_ is the time-varying effective reproduction number and *f*_*i*_ is the infectivity profile. The infectivity profile is a probability mass function that describes the relative contributions of infected individuals in the different compartments to overall transmission. We model the infectivity profile *f*_*i*_ using a modified negative binomial distribution. Our modification of the negative binomial distribution involves setting the probability mass in compartment *i* = 0 to 0 (that is, *f*_*i*_ = 0) and renormalizing the overall probability mass to 1. Cumulative case incidence is given by:

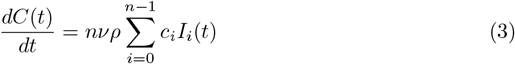

where *ρ* is the overall reporting rate (that is, the probability that an infected individual gets reported as a case) and *c*_*i*_ is the case detection profile. The case detection profile is a probability mass function that describes the relative probabilities of a reported case stemming from the various *I* compartments. Like *f*_*i*_, we model *c*_*i*_ using a modified (*c*_0_ = 0) and renormalized negative binomial distribution. Case incidence over a period of time (for example, weekly case incidence) is calculated using *C*, specifically by taking the difference between cumulative case incidence at the later timepoint *t*_*l*_ and cumulative case incidence at the earlier timepoint *t*_*e*_: *C*(*t*_*l*_) − *C*(*t*_*e*_). Wastewater virus concentrations are modeled as follows:

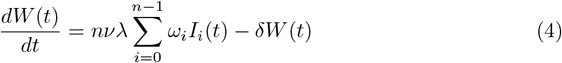

where *λ* is the shedding load scaling constant, *ω*_*i*_ is the shedding load profile, and *δ* is the viral outflow rate from the wastewater. The shedding load scaling constant *λ* can be interpreted as the total amount of virus shed by an average individual into the wastewater system over the course of their infection. The shedding load profile is a probability mass function that describes the relative contributions of viral shedding from individuals in each of the various infection compartments. Like *f*_*i*_ and *c*_*i*_, we model *ω*_*i*_ using a modified (*ω*_0_ = 0) and renormalized negative binomial distribution.

We forward simulate this process model using the following Euler-Maruyama set of equations:

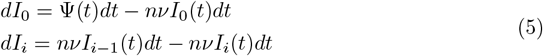

where *dt* is the size of the time step. The *I* state variables are updated as follows:

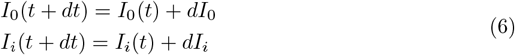

Similar to the approaches taken by Goldstein et al. (2023) and Watson et al. (2024), we model the time-varying effective reproduction number *R*_*t*_ as a separate variable in the process model. This has been traditionally done in the field of quantitative genetics where a phenotypic trait can evolve over time and impact population state variables.

Specifically, and as proposed in Cazelles et al. (2018), we use a Brownian motion model to model changes in *R*_*t*_ over time:

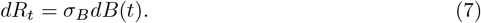

Here, *dB*(*t*) are independent and identically distributed normal random variables with mean 0 and variance *dt* (Butler and King, 2004). The parameter *σ*_*B*_ quantifies the strength of Brownian motion. The effective reproduction number is then updated similarly to the other state variables:

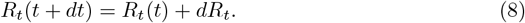

Given the range of possible values that *R*_*t*_ can take on, we reset *R*_*t*_(*t* + *dt*) to zero if negative. Cumulative incidence and wastewater virus concentrations are similarly forward-simulated using Euler-Maruyama equations:

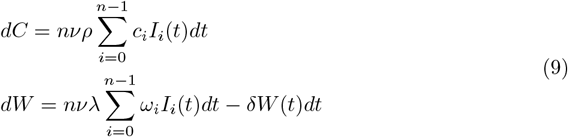

where *dt* is the size of the time step. Cumulative incidence and wastewater virus concentration state variables are updated as follows:

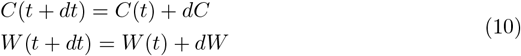

#### 2.1.2 The observation models

The observation models relate the observed data to the state variables *C* and *W*. For case data, we use a negative binomial distribution, where the probability of observing *N*_*c*_ cases over a time period from *t*_*e*_ to *t*_*l*_ is given by:

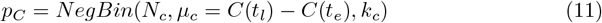

where early and late timepoints are denoted *t*_*e*_ and *t*_*l*_, respectively, *µ*_*c*_ denotes the mean of the negative binomial distribution and *k*_*c*_ denotes the distribution’s overdispersion parameter. For wastewater virus concentration data, we use a gamma distribution as our observation model, where the probability of observing virus concentration *N*_*w*_ is given by:

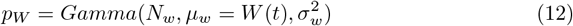

where *µ*_*w*_ denotes the mean of the gamma distribution and 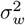denotes the distribution’s variance.

### 2.2 The bootstrap particle filter

We use a bootstrap particle filter (Gordon et al., 1993) to estimate model parameters and reconstruct latent state variables, most notably the time-varying effective reproduction number *R*_*t*_. When interfacing the model with case incidence data, the likelihood of each observed case incidence count is calculated using equation (11). When interfacing the model with wastewater concentration data, the likelihood of each observed wastewater measurement is calculated using equation (12). For joint estimation, when both case incidence and wastewater data are available, we assume independence and obtain the joint probability by multiplying the individual likelihoods. At time points when only case incidence data or only virus concentration data are available, we use the equation relevant to the observed data point.

Particles are simulated until they are resampled at each observation time using a multinomial sampling scheme. Resampling is performed with replacement and based on particle weights, which are given by their calculated likelihoods (that is, for each particle, the probability of observing the data point(s) given the model prediction generated by the particle). Resampled particles are then propagated until the next observation point. The latent state variables are reconstructed by randomly drawing one particle from the set present at the last observation time point. This gives us a trajectory of reconstructed *R*_*t*_ values, as well as reconstructed values of cumulative case data *C* and wastewater concentrations *W*. The marginal likelihood (or marginal log-likelihood), under a given model parameterization, is computed as follows. We first calculate the arithmetic mean of the particle weights at each observation time *t*:

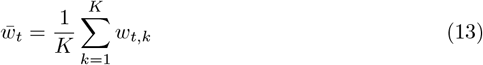

where *k* denotes the individual particles, *K* is the total number of particles, and *w*_*t,k*_ is the weight of particle *k* at time *t*. We then take the product of these mean weights across observation times. The marginal log-likelihood is then computed by summing the log mean particle weights across observation time points:

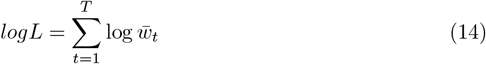

We calculate the marginal log-likelihoods using observation time points that occur after a specified burn-in period (*t > t*_*burn−in*_).

### 2.3 Simulation of the mock datasets

To aid in the development and testing of our statistical approach for estimating *R*_*t*_, we generated two mock datasets: one that comprises time series of reported cases and another corresponding one that comprises time series of wastewater virus concentrations. We let the process model for simulating the mock datasets contain *n* = 40 *I* compartments and set the average duration of time an individual spent in an infectious compartment to 1*/*(*nν*) = 1 day. On average, an individual therefore spent 40 days from the time of infection until leaving the last *I* class. As such, the model allows for an individual to transmit infection, be detected as a case, and shed into the wastewater for up to approximately 40 days.

The infectivity profile (*f*_*i*_), case detection profile (*c*_*i*_), and shedding load profile (*ω*_*i*_) we used to simulate the mock datasets are shown in Figures 1A-C. These profiles were parameterized to recapitulate time-since-infection distributions of infectivity, case detection, and shedding, respectively that have been previously used in the literature based on empirical data (Huisman et al., 2022) (Figures 1D-F). Specifically, we chose parameter values for the *f*_*i*_, *c*_*i*_ and *ω*_*i*_ profiles by drawing 10,000 samples from the time-since-infection distributions assumed in Huisman et al. (2022) and finding the parameter values of the modified negative binomial distributions that yield the highest likelihoods based on these samples. We further set the reporting rate to *ρ* = 0.1, the shedding load constant to *λ* = 5000 genome copies/ml, the virus outflow rate to *δ* = 0.5 genome copies/mL/day (corresponding to a virus half-life of 2 days in wastewater).

**Figure 1.**
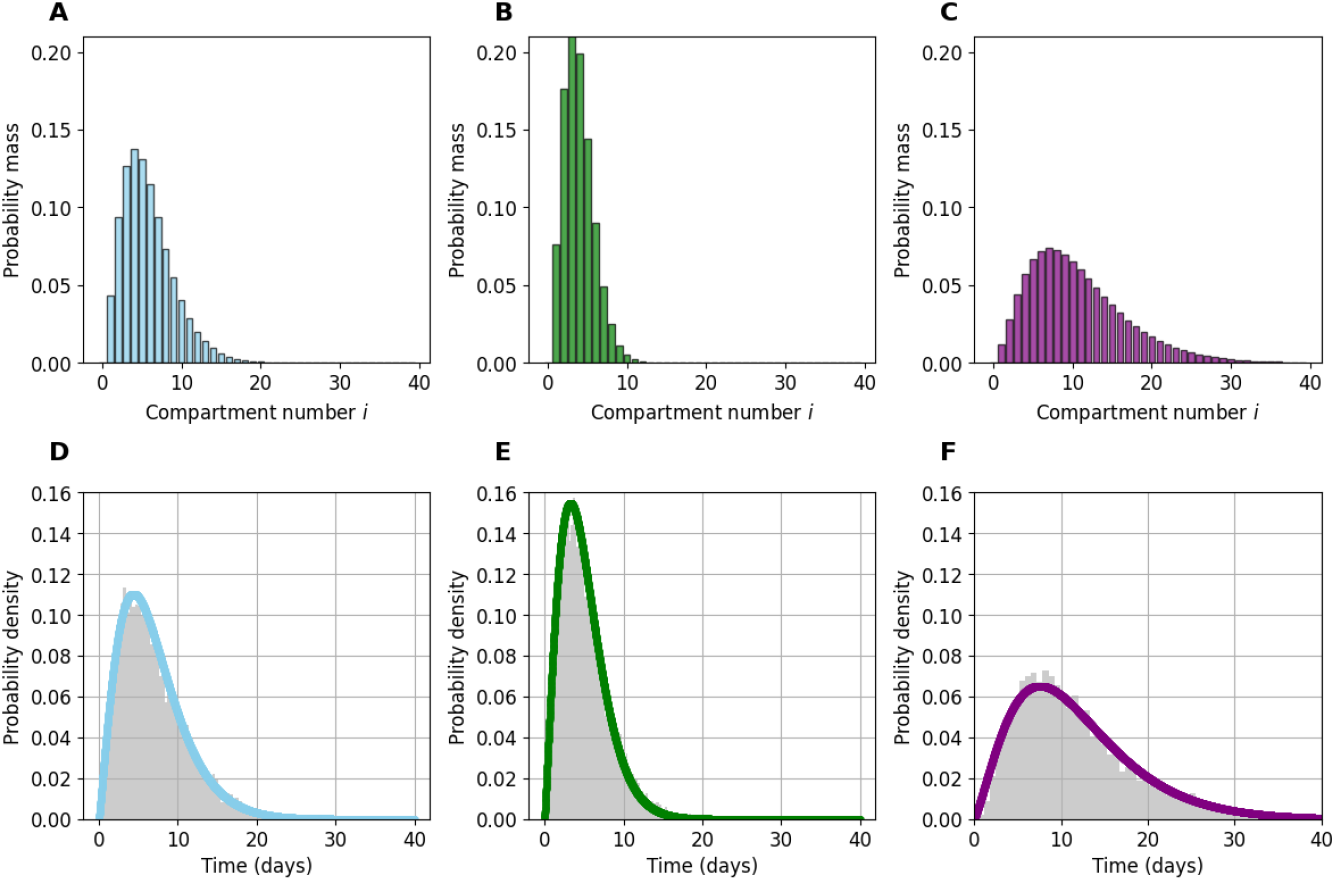
Parameterization of infectivity, case detection, and shedding load profiles for the mock dataset. (A) The infectivity profile *f*_*i*_, given by a modified negative binomial distribution with parameters *r* = 3.90 and *p* = 0.45. (B) The case detection profile *c*_*i*_, given by a modified negative binomial distribution with parameters *r* = 11.53 and *p* = 0.80. (C) The shedding load profile *ω*_*i*_, given by a modified negative binomial distribution with parameters *r* = 3.15 and *p* = 0.24. We model a total of *n* = 40 compartments, indexed from *i* = 0 to *i* = *n −* 1. The negative binomial distributions in panels (A)-(C) are modified by setting *f*_0_, *c*_0_, and *ω*_0_ to 0, respectively, and renormalizing the probability masses to sum to 1. (D) Infectivity profile shown as a function of time since infection. (E) Case detection profile shown as a function of time since infection. (F) Shedding load profile shown as a function of time since infection. Gray distributions in panels (D)-(F) show 10000 samples from the time-since-infection distributions assumed in Huisman et al. (2022).

Instead of using the Brownian motion model to simulate *R*_*t*_ (equations (7) and (8)), we parameterized *R*_*t*_ using a concatenated combination of sine functions of various periods and amplitudes. Finally, we set the initial conditions of *I*_*i*_, for 0 ≤ *i* ≤ (*n*− 1) to 1 for all *i* and simulated the model for 10 years. From this forward simulation, we extracted a two-year period that by eye appeared dynamically interesting, with infection waves that lasted on the order of a couple of months. The time-varying effective reproduction number *R*_*t*_ for this extracted period of time is shown in Figure 2A. The number of individuals in infection classes *i* = 0 through *i* = *n* −1 over this same time period is shown in Figure 2B.

**Figure 2.**
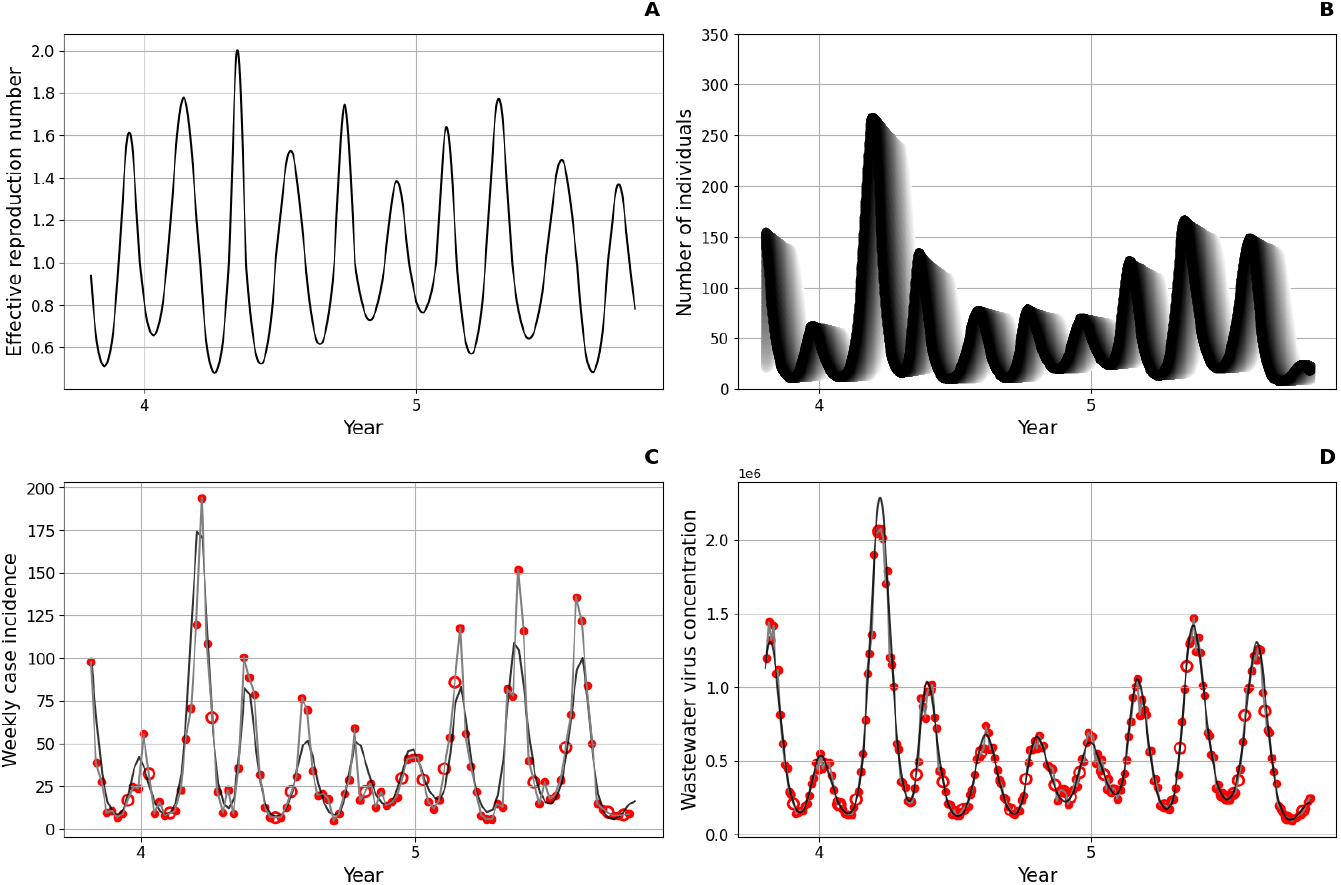
Dynamics of the mock datasets. (A) The time-varying effective reproduction number *R*_*t*_. (B) The dynamics of infected individuals *I*_*i*_. The dynamics of *I*_0_ are shown in dark black. The dynamics of higher-*i* infection classes are shown in a progression to grayer shades. (C) Weekly case incidence. The black line shows ‘true’ simulated case incidence. Red filled circles show case incidence with added measurement noise. 10% of the weekly case data are removed at random and considered as missing. Missing data are shown with red unfilled circles. A gray line connects measured case incidence values. (D) Virus concentrations in wastewater (in genome copies/mL). Black line shows true concentrations *W* (*t*). Red filled circles show virus concentrations sampled every 3 days with added measurement noise. 10% of the wastewater data points are removed at random and considered as missing. Missing data are shown with red unfilled circles. A gray line connects measured virus concentration data.

Weekly cases were calculated from the simulated cumulative case count variable *C*. To generate the mock dataset of weekly cases, we further added measurement noise to these ‘true’ weekly case counts. Measurement noise was added to each weekly case incidence count using a negative binomial distribution, with the mean given by the weekly case count and an overdispersion parameter of *k*_*c*_ = 10. To reflect incomplete data that may be encountered in empirical datasets, we further implemented data loss by letting each data point have only a 90% probability of being recorded. ‘True’ weekly case counts as well as the case counts with additional measurement noise and data loss are shown in Figure 2C.

Wastewater samples were assumed to be taken every 3 days. Measurement noise was added to the simulated wastewater virus concentrations *W* using a gamma distribution with a mean given by the simulated virus concentration and a coefficient of variation of 0.10. This assumption leads to a constant shape parameter for the gamma distribution of *α* = 100 and a time-varying scale parameter for the gamma distribution of *θ*(*t*) = *W* (*t*)*/α*. To again reflect incomplete data that may be encountered in empirical datasets, we further implemented data loss by letting each data point have only a 90% probability of being recorded. ‘True’ wastewater virus concentrations as well as the virus concentrations with additional measurement noise and data loss are shown in Figure 2D.

Our mock datasets to develop and test our statistical approach to estimate *R*_*t*_ comprise the simulated noisy case data (shown in red) and the simulated noisy wastewater data (shown in red) in Figures 2C and 2D, respectively.

### 2.4 Application of the bootstrap particle filter to the mock datasets

We used the bootstrap particle filter described in Section 2.2 to estimate a subset of the parameters of the state-space model described in Section 2.1, while keeping other parameters fixed at given values. Specifically, we set the number of *I* compartments to *n* = 40 and set the overall average duration of being in the infectious classes as 1*/ν* = 40 days. We further set the infectivity profile *f*_*i*_, the case detection profile *c*_*i*_, and the shedding load profile *ω*_*i*_ to their true values (that is, to the values used in the forward simulation of the mock datasets; Figures 1A-C) and the viral outflow rate *δ* to its true value of 0.5 genome copies/mL/day. When interfacing the model with only case data, we also set the overdispersion parameter *k*_*c*_ of the case observation model to its true value of 10 and sought to estimate the reporting rate *ρ* and the Brownian motion parameter *σ*_*B*_. When interfacing the model with only wastewater data, we also set the coefficient of variation (*σ*_*w*_*/µ*_*w*_) of the wastewater observation model to its true value of 0.10 and sought to estimate the shedding load scaling constant *λ* and the Brownian motion parameter *σ*_*B*_. Finally, when interfacing the model with both case data and wastewater data, we attempted to jointly estimate *ρ* and *λ*. In all of our analyses, we also reconstructed the time-varying reproduction number *R*_*t*_.

In the bootstrap particle filter, we set the number of particles to 1000. For each particle, the initial number of individuals in the *I*_0_ compartment was drawn from a lognormal (*log*_10_) distribution with mean 2.5 and standard deviation of 1. The remaining *I*_*i*_ compartments were initialized to 0. For each particle, the initial *R*_*t*_ value was also drawn from a lognormal (*log*_10_) distribution, with a mean of 0 and a standard deviation of 0.5. Particle weights were calculated as described in Section 2.2, and the marginal log-likelihood was calculated by summing the logs of the mean particle weights over all observation time points following a burn-in period of 70 days.

### 2.5 Application of the bootstrap particle filter to the Zurich SARS-CoV-2 datasets

To demonstrate our approach on an empirical dataset, we used existing SARS-CoV-2 datasets from Zurich that were originally analyzed in Huisman et al. (2022) and made publicly available by the authors on a GitHub site. The case incidence data are reported on a daily basis and span the time period February 26, 2020 through April 7, 2021, with data missing for a total of 20 days throughout this period. A 7-day periodicity is apparent in this case dataset (Figure 3A), indicating that reporting rates likely differed by day of week. The wastewater data derive from samples collected over the time period September 3, 2020 through January 19, 2021 (Figure 3B), with measurements taken twice a week (on Thursdays and Sundays). These data consist of measured RNA loads from two assays: one that targeted the N1 region of the nucleocapsid protein (N) gene and one that targeted the N2 region of this gene. Measured RNA loads targeting these regions were strongly correlated (Figure 3B). Measurements are missing for a total of 51 days throughout this period, predominantly in the months of September and October 2020. In our statistical analyses, we excluded three time points from the wastewater data: December 13, 2020, January 10, 2021, and January 12, 2021. The December 13, 2020 and January 12, 2021 time points were removed because of discrepancies between the N1 and N2 measurements, where virus concentrations measured by these markers differed from one another by approximately 35% and 50%, respectively, when all other N1 and N2 measurements fell within 25% of one another. The January 10, 2021 time point was removed because it was a major outlier, exceeding temporally adjacent timepoints by over 160%. Because of the strong correlation between the N1 and N2 datasets, in our analyses, we used only the N1 dataset. Because we aimed to test our inference approach on case data, on wastewater data, and on a combination of these two datasets, we truncated both datasets to the same time period of observations between September 3, 2020 and January 19, 2021.

**Figure 3.**
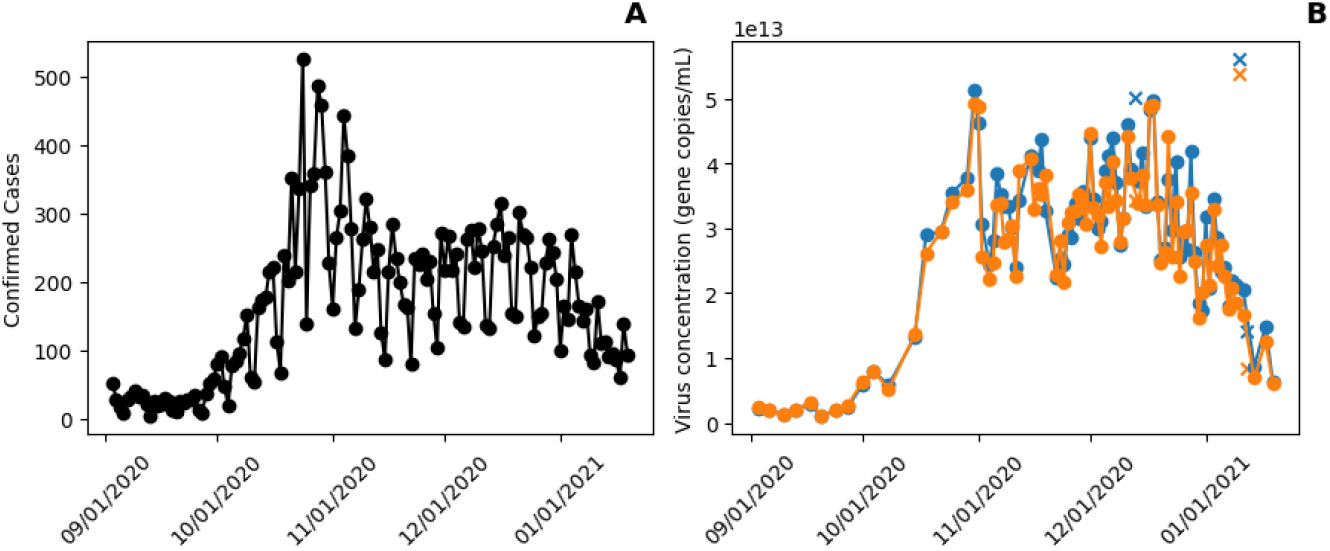
SARS-CoV-2 data from Zurich, Switzerland that were used to test our particle filtering inference approach. (A) Daily case incidence data. (B) Wastewater virus concentration data. RNA viral loads measured using the N1 assay are shown in blue. RNA viral loads measured using the N2 assay are shown in orange. Each of the three excluded time points are marked with an *×* on both the N1 and N2 data. The case data and the wastewater data were truncated to span the period September 3, 2020 through January 19, 2021.

We again used the bootstrap particle filter described in Section 2.2 to estimate a subset of the parameters of the state-space model, while keeping other parameters fixed at given values. We again set the number of *I* compartments to *n* = 40 and set the overall average duration of being in the infectious classes as 1*/ν* = 40 days. We again set the infectivity profile *f*_*i*_, the case detection profile *c*_*i*_, and the shedding load profile *ω*_*i*_ to the profiles shown in Figures 1A-C and set the viral outflow rate *δ* this time to 1 gene copies/mL/day. When interfacing the model with only case data, we sought to estimate the reporting rate *ρ* and *σ*_*B*_. To accommodate the day-of-week periodicity apparent in Figure 3A, we identified consecutive four-day time windows corresponding to Friday, Saturday, Sunday, and Monday. For each identified 4-day time window, we computed the ratio of weekend case incidence (sum of Saturday and Sunday incidence) to weekday case incidence (sum of Friday and Monday incidence). The median of these calculated ratios over all available four-day windows was 0.533. We took this median value as the reporting rate adjustment factor. In applying the bootstrap particle filter to the Zurich case data, this weekend adjustment factor was incorporated by discounting the reporting rate *ρ* on weekends by this factor to allow the model to better capture the weekly periodicity in the observed case data. When interfacing the model with only wastewater data, we sought to estimate the shedding load scaling constant *λ* and *σ*_*B*_. Finally, when interfacing the model with both case data and wastewater data, we attempted to jointly estimate *ρ* and *λ*, while setting *σ*_*B*_ to a reasonable value based on our analyses with just the case incidence data and just the wastewater data.

For the case observation model, we set the overdispersion parameter *k*_*c*_ to a value of 10, reflecting little overdispersion. For the wastewater observation model, we decided to parameterize observation noise with a constant standard deviation, using the N1 and N2 data to empirically estimate this standard deviation. To approximate this standard deviation, we fit a Gaussian distribution with mean zero to the difference in the N1 and N2 measurements. This yielded an estimate of *σ* = 2.2114 *×* 10^12^ gene copies/mL, which we used to parameterize the gamma distribution shown in equation (12). Specifically, we set the shape parameter *α* to 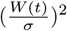 and the scale parameter *θ* to 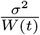. With this parameterization, the resulting gamma distribution has a mean given by the simulated virus concentration *W* (*t*) and a constant variance of *σ*^2^.

In the bootstrap particle filter, we again set the number of particles to 1000. For each particle, the initial number of individuals in the *I*_0_ compartment was drawn from a lognormal (*log*_10_) distribution with mean 2.5 and standard deviation of 1. The remaining *I*_*i*_ compartments were initialized to 0. For each particle, the initial *R*_*t*_ value was drawn from a lognormal (*log*_10_) distribution with a mean of 0 and a standard deviation of 0.5. Particle weights were again calculated as described in Section 2.2, and the marginal log-likelihood was calculated by summing the logs of the mean particle weights over all observation time points following a burn-in period of 18 days.

Based on our results (below), we additionally considered a modified process model with an additional stochastic component. Specifically, we allowed for environmental stochasticity in the viral outflow rate *δ* in impacting viral concentration levels in wastewater:

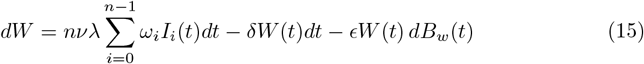

where the parameter *ϵ* quantifies the strength of environmental noise impacting viral decay dynamics and *dB*_*w*_(*t*) are independent and identically distributed normal random variables: *dB*_*w*_(*t*) ∼ 𝒩 (0, *dt*) (Butler and King, 2004).

Finally, a study by Emmenegger et al. (2023) made available serological data for Zurich. This study investigated the temporal evolution of SARS-CoV-2 seropositivity in two independent cohorts: patients at the University Hospital of Zurich (USZ) and healthy blood donors who donated at the Blood Donation Service of the Canton of Zurich (BDS). Seroprevalence in both of these cohorts remained low during the summer months of 2020 before increasing sharply in October 2020. We used the seroprevalence estimates from the BDS cohort in mid-September 2020 (0.8% [95% CI: 0.5-1.2]) and mid-December 2020 (5.1% [95% CI: 4.2–6.4]) in our analyses below. This study reported mid-December 2020 values for both cohorts, with the USZ cohort arriving at similar seropositivity estimates (6.3% [95% CI: 5.5–7.2]). The mid-September 2020 values were only plotted graphically and we therefore used the PlotDigitizer website to extract the data for this earlier time point seropositivity estimate. Between mid-September and mid-December 2020, the increase in seropositivity was therefore approximately 4.3%. To incorporate seropositivity as a variable in our process model, we assumed a delay of approximately 13 days on average from the time of infection to becoming seropositive, based on a SARS-CoV-2 study that estimated a period of 11-14 days between infection and becoming seropositive (Kellam and Barclay, 2020). With this assumption, we modeled the increase in the number of individuals who are seropositive to SARS-CoV-2 using the equation:

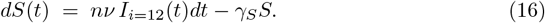

where the first term captures increases in seropositivity at the level of the population and the second term captures the loss of seropositivity that occurs either through seropositive individuals leaving the population (through mortality or emigration) or through seroreversion through antibody waning. Because the time period of the Zurich data spanned only 3-4 months, we assumed that the loss of seropositivity over this time frame was negligible and set *γ*_*S*_ to 0 in our analyses. The initial number of seropositive individuals was arbitrarily set to *S*(0) = 0. The number of individuals who seroconverted between mid-September and mid-December was calculated as the difference between the value of *S* on December 16, 2020 and the value of *S* on September 15, 2020. This number was divided by the canton of Zurich’s population size to get simulated estimates of the change in seropositivity over this time period. The total population was assumed to be 1,553,423 individuals (Swiss Federal Statistical Office, 2020).

### 2.6 Forecasting using the bootstrap particle filter

To forecast *R*_*t*_, case incidence, and wastewater virus concentrations, we forward simulated the state space model for an additional 10 days. To visualize these forecasts, we randomly chose 10 of the 1000 particles to plot.

## 3 Results

### 3.1 *R*_*t*_ Estimation from the Mock Datasets

#### 3.1.1 *R*_*t*_ Estimation from the Mock Case Data

Using only the mock weekly case incidence data (Figure 2C), we first asked whether the bootstrap particle filter could recover the ‘true’ *R*_*t*_ trajectory shown in Figure 2A. Towards this end, we ran the bootstrap particle filter, parameterized using a range of different reporting rates (*ρ* values) and a range of different Brownian motion strengths (*σ*_*B*_ values). The resulting log-likelihood surface is shown in Figure 4A. There is a vertical band (ridge) of highest-likelihood solutions, with the value of *σ*_*B*_ = 0.66 yielding the highest likelihoods. Interesting, the value of *ρ* does not appear to impact the likelihoods. To better understand why this is the case, we simulated the particle filter with a value of *σ*_*B*_ = 0.66 and using two different reporting rates: *ρ* = 0.2 and *ρ* = 0.8. Both reporting rate values yield quantitatively similar *R*_*t*_ trajectories (Figure 4B) and similar reconstructed weekly case incidence (Figure 4C). Reconstructed infection prevalence patterns (values of *I*_*i*_ for 0 ≤ *I* ≤ (*n* − 1)) for these two values of *ρ*, however, differ substantially from one another (Figure 4D). The low reporting rate of *ρ* = 0.2 results in higher infection prevalence than the higher reporting rate of *ρ* = 0.8. These results, along with inspection of equation (3), indicates that *ρ* is unidentifiable within the term ∑*ρI*_*i*_(*t*). Therefore, and consistent with earlier findings (Watson et al., 2024), we cannot accurately reconstruct underlying infection dynamics or infer reporting rates in the absence of additional data sources.

**Figure 4.**
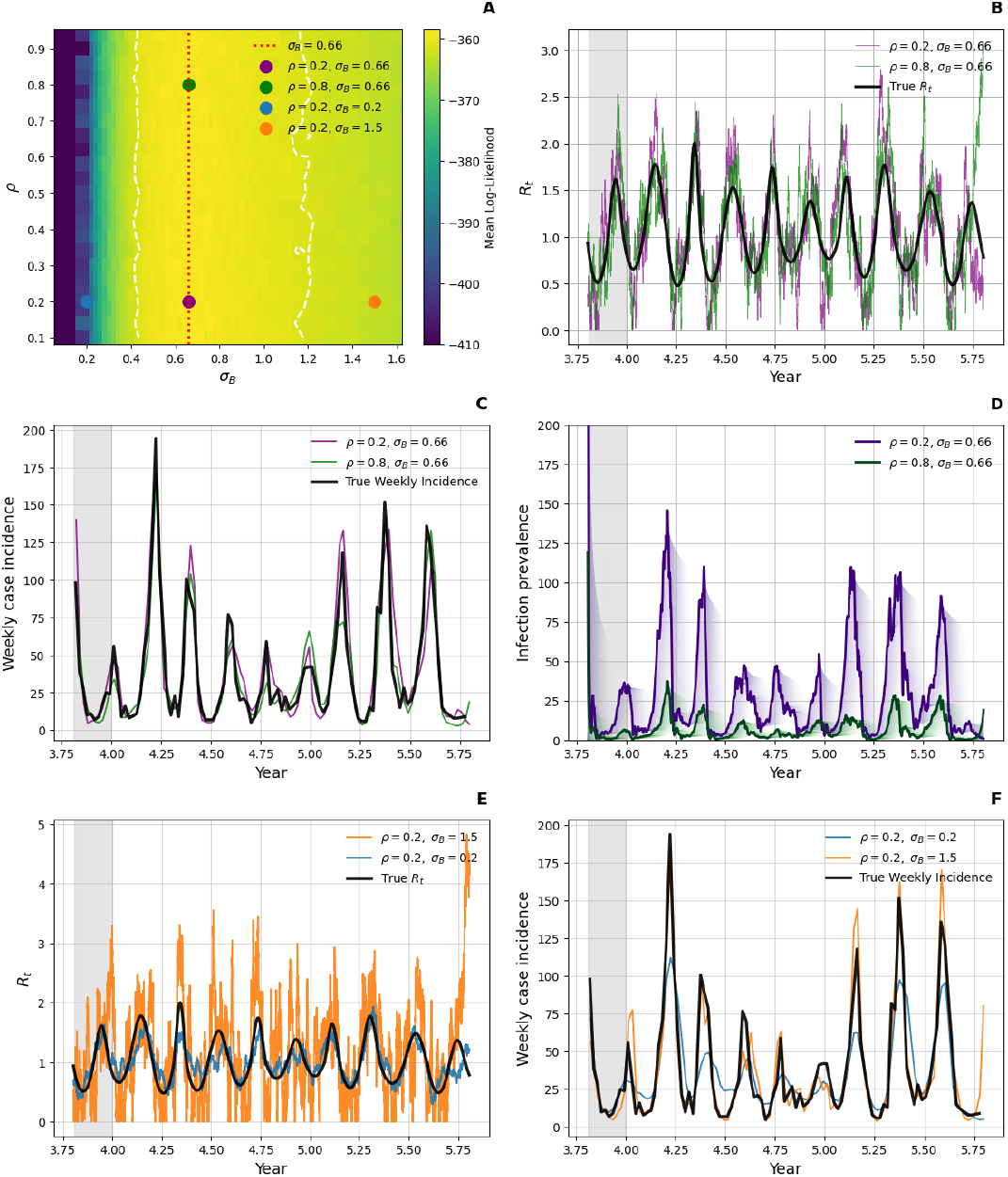
Estimation of the reporting rate *ρ* and the strength of Brown motion *σ*_*B*_ from the mock case dataset. (A) Heatmap of log-likelihood values. Values below −410 were set to −410 for improved visualization. The red dash line indicates the maximum log-likelihood of *σ*_*B*_. The lower and upper 95% confidence intervals are shown with dashed white lines. The purple and green dots mark two (*ρ, σ*_*B*_) parameter combinations that lie along the red dashed line. The blue, purple and orange dots mark three (*ρ, σ*_*B*_) parameter combinations that lie along the *ρ* = 0.2 line. We considered the first 70 days as a burn-in period and as such removed this period from the log-likelihood calculations. At each parameter combination, the log-likelihood value shown is the median value across 10 bootstrap particle filters. (B) Reconstructions of the time-varying effective reproduction number *R*_*t*_ under the two different (*ρ, σ*_*B*_) parameter combinations shown with purple and green dots in panel A. True *R*_*t*_ values used to generate the mock data are shown alongside these reconstructed values. (C) Reconstructions of weekly case incidence under the two different parameter combinations of (*ρ* = 0.2, *σ*_*B*_ = 0.66) and (*ρ* = 0.8, *σ*_*B*_ = 0.66), alongside the mock weekly case incidence data. (D) Reconstructions of infection prevalence under the two different parameter combinations of (*ρ* = 0.2, *σ*_*B*_ = 0.66) and (*ρ* = 0.8, *σ*_*B*_ = 0.66). The dynamics of *I*_0_ are shown in bold. The dynamics of higher-*i* infection classes are shown in lighter shades. (E) Reconstructions of the time-varying effective reproduction number *R*_*t*_ under the two different (*ρ, σ*_*B*_) parameter combinations shown with blue and orange dots in panel A. True *R*_*t*_ values used to generate the mock data are shown alongside these reconstructed values. (F) Reconstructions of weekly case incidence under the two different parameter combinations of (*ρ* = 0.2, *σ*_*B*_ = 0.2) and (*ρ* = 0.2, *σ*_*B*_ = 1.5), alongside the mock weekly case incidence data. In panels B-F, the 70-day burn-in period is shown with a gray band.

Finally, we looked at the reconstructed *R*_*t*_ trajectories and weekly case incidence for two different values of *σ*_*B*_ (0.20 and 1.50) while fixing the reporting rate at *ρ* = 0.2 (Figure 4E,F). A *σ*_*B*_ of 0.20 fell below the optimal value of *σ*_*B*_ = 0.66. A *σ*_*B*_ of 1.5 fell above the optimal value of *σ*_*B*_ = 0.66. When *σ*_*B*_ = 0.2, the reconstructed *R*_*t*_ trajectory fails to capture the higher amplitude peaks of the true *R*_*t*_ fluctuations (Figure 4E) and reconstructed weekly case incidence similarly fails to capture the observed peaks and troughs in the mock case data (Figure 4F). When *σ*_*B*_ = 1.5, the high variability in *R*_*t*_ causes the *R*_*t*_ reconstruction to be excessively jagged (Figure 4E), although reconstructions of weekly case incidence do not deviate substantially from those observed from the mock data set (Figure 4F). Our intermediate value of *σ*_*B*_ = 0.66 effectively captures the amplitude of the true *R*_*t*_ fluctuations without introducing the excessive jaggedness observed at higher *σ*_*B*_ values (Figure 4B) and, as indicated by the log-likelihood surface (Figure 4A), does account better for the observed case data. These results indicate that the case data do indeed allow us to ‘choose’ an appropriate *σ*_*B*_ value that manages to reproduce the true fluctuations in *R*_*t*_ and to faithfully reconstruct weekly case incidence.

#### 3.1.2 *R*_*t*_ Estimation from the Mock Wastewater Data

Next, using only the mock wastewater data (Figure 2D), we asked whether the bootstrap particle filter could recover the ‘true’ *R*_*t*_ trajectory shown in Figure 2A. Towards this end, we again ran the bootstrap particle filter, parameterized using a range of different shedding load constants (*λ* values) and a range of different Brownian motion strengths (*σ*_*B*_ values). The resulting log-likelihood surface is shown in Figure 5A. Again, there is a ridge of highest-likelihood solutions, with the value of *σ*_*B*_ = 0.56 this time yielding the highest likelihoods. The value of *λ* does not appear to impact the likelihoods. To better understand why this is the case, we simulated the particle filter with a value of *σ*_*B*_ = 0.56 and using two different *λ* values: *λ* = 8, 000 and *λ* = 14, 000 copies/mL. Both *λ* values yield quantitatively similar *R*_*t*_ trajectories (Figure 5B) and similar reconstructed wastewater virus concentration trajectories (Figure 5C). Reconstructed infection prevalence patterns (values of *I*_*i*_ for 0 ≤ *i*≤ (*n*− 1)) for these two values of *λ*, however, again differed substantially from one another (Figure 5D). The low shedding constant of *λ* = 8, 000 copies/mL results in higher infection prevalence than the higher shedding constant of *λ* = 14, 000 copies/mL. These results, along with inspection of equation (4), indicates that *λ* is unidentifiable within the term ∑*λI*_*i*_(*t*). Therefore, and again consistent with earlier findings (Watson et al., 2024), we cannot accurately reconstruct underlying infection dynamics or infer the shedding load constant in the absence of additional data.

**Figure 5.**
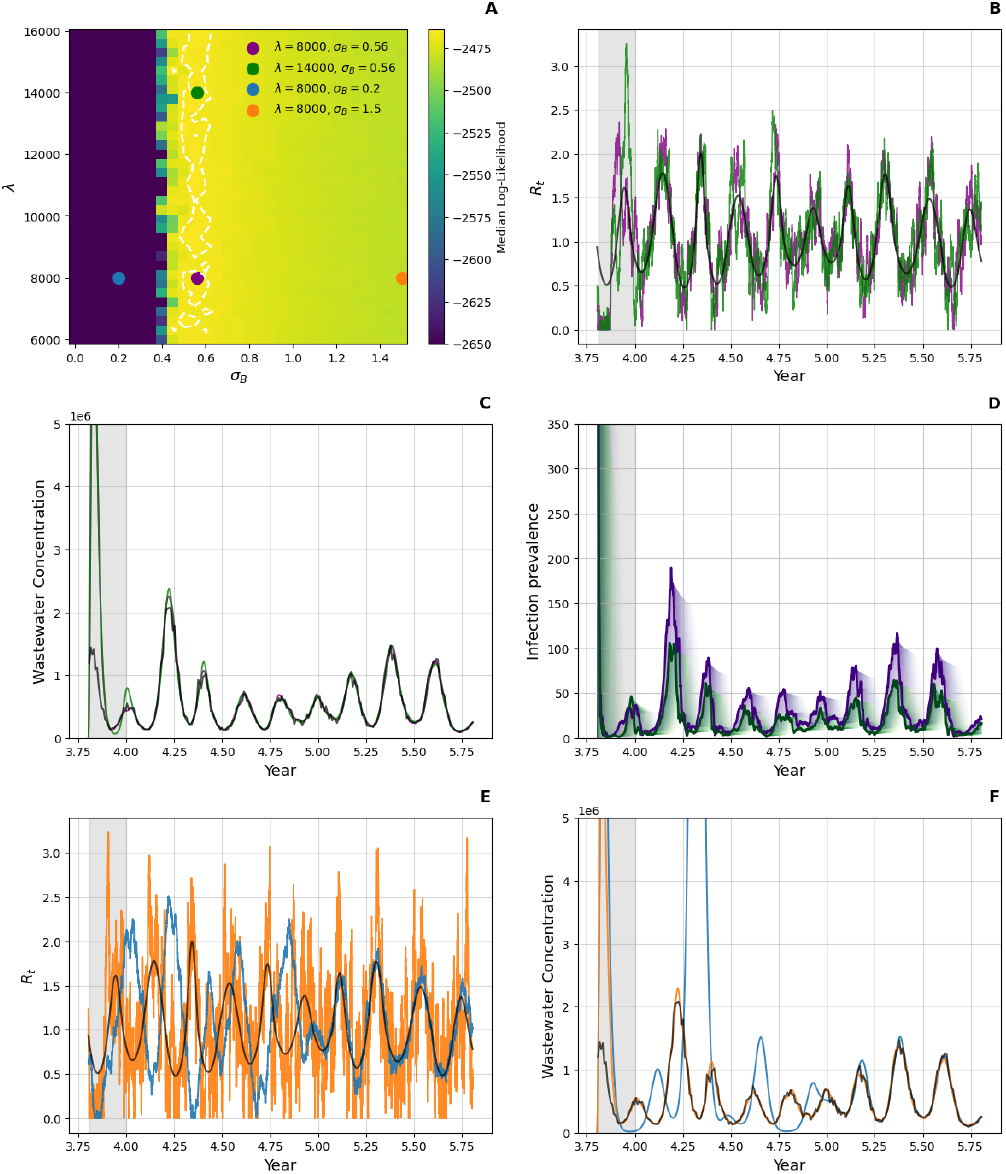
Estimation of the shedding load constant *λ* and the strength of Brown motion *σ*_*B*_ from the mock wastewater dataset. (A) Heatmap of log-likelihood values. Values below −2650 were set to −2650 for improved visualization. The red dash line indicates the maximum log-likelihood of *σ*_*B*_. The lower and upper 95% confidence intervals are shown with dashed white lines. The green and purple dots mark two (*λ, σ*_*B*_) parameter combinations that lie along the red dashed line. The blue, purple and orange dots mark three (*λ, σ*_*B*_) parameter combinations that lie along the *λ* = 8000 copies/mL line. We considered the first 70 days as a burn-in period and as such removed this period from the log-likelihood calculations. At each parameter combination, the log-likelihood value shown is the median value across 10 bootstrap particle filters. (B) Reconstructions of the time-varying effective reproduction number *R*_*t*_ under the two different (*λ, σ*_*B*_) parameter combinations shown with purple and green dots in panel A. True *R*_*t*_ values used to generate the mock data are shown alongside these reconstructed values. (C) Reconstructions of wastewater concentration under the two different parameter combinations of (*λ* = 8000, *σ*_*B*_ = 0.56) and (*λ* = 14, 000, *σ*_*B*_ = 0.56), alongside the mock wastewater concentration data. (D) Reconstructions of infection prevalence under the two different combinations of (*λ* = 8000, *σ*_*B*_ = 0.56) and (*λ* = 14, 000, *σ*_*B*_ = 0.56). The dynamics of *I*_0_ are shown in bold. The dynamics of higher- *i* infection classes are shown in lighter shades. (E) Reconstructions of the time-varying effective reproduction number *R*_*t*_ under the two different (*λ, σ*_*B*_) parameter combinations shown with blue and orange dots in panel A. True *R*_*t*_ values used to generate the mock data are shown alongside these reconstructed values. (F) Reconstructions of wastewater virus concentrations under the two different parameter combinations of (*λ* = 8000, *σ*_*B*_ = 0.2) and (*λ* = 8000, *σ*_*B*_ = 1.5), alongside the mock wastewater virus concentration data. In panels B-F, the 70-day burn-in period is shown with a gray band.

Finally, we looked at the reconstructed *R*_*t*_ trajectories and wastewater virus concentration data for two different values of *σ*_*B*_ (0.20 and 1.50) while fixing *λ* at 8000 copies/mL. When *σ*_*B*_ = 0.2, the reconstructed *R*_*t*_ trajectory again fails to capture the higher amplitude peaks of the true *R*_*t*_ fluctuations (Figure 5E) and reconstructed wastewater concentrations similarly fail to capture the observed peaks and troughs in the mock wastewater data (Figure 5F). When *σ*_*B*_ = 1.50, the high variability in *R*_*t*_ causes the *R*_*t*_ reconstruction to again be excessively jagged (Figure 5E), although, again, reconstructions of the wastewater virus concentrations do not deviate substantially from those observed in the mock data set (Figure 5F). Our intermediate value of *σ*_*B*_ = 0.56 effectively captures the amplitude of the true *R*_*t*_ fluctuations (Figure 5B) and better accounts for the observed wastewater data (Figure 5A,B).These results indicate that the wastewater virus concentration data do indeed allow us to ‘choose’ an appropriate *σ*_*B*_ value that manages to reproduce the true fluctuations in *R*_*t*_ and to faithfully reconstruct wastewater virus concentrations.

#### 3.1.3 *R*_*t*_ Estimation from Both Mock Case Data and Mock Wastewater Data

We now turn to assessing the potential for jointly estimating *ρ* and *λ* when both case incidence and wastewater virus concentration levels are available as data streams. We again used the same model structure as before, fixing the profiles *f*_*i*_, *c*_*i*_, and *ω*_*i*_ to their true values. The strength of Brownian motion *σ*_*B*_ were set to 0.61, corresponding to the average of the case-derived and wastewater-derived estimates of *σ*_*B*_ of 0.66 and 0.56, respectively. We ran the bootstrap particle filter across a range of reporting rates *ρ* and across a range of shedding load constants *λ* to determine how combinations of *ρ* and *λ* impacted model likelihoods. Figure 6A shows the resulting log-likelihood surface. A high-likelihood ridge is apparent, spanning from low-*ρ*, low-*λ* parameter combinations to high-*ρ*, high-*λ* parameter combinations. This indicates that many different (*ρ, λ*) combinations can produce comparable likelihoods.

**Figure 6.**
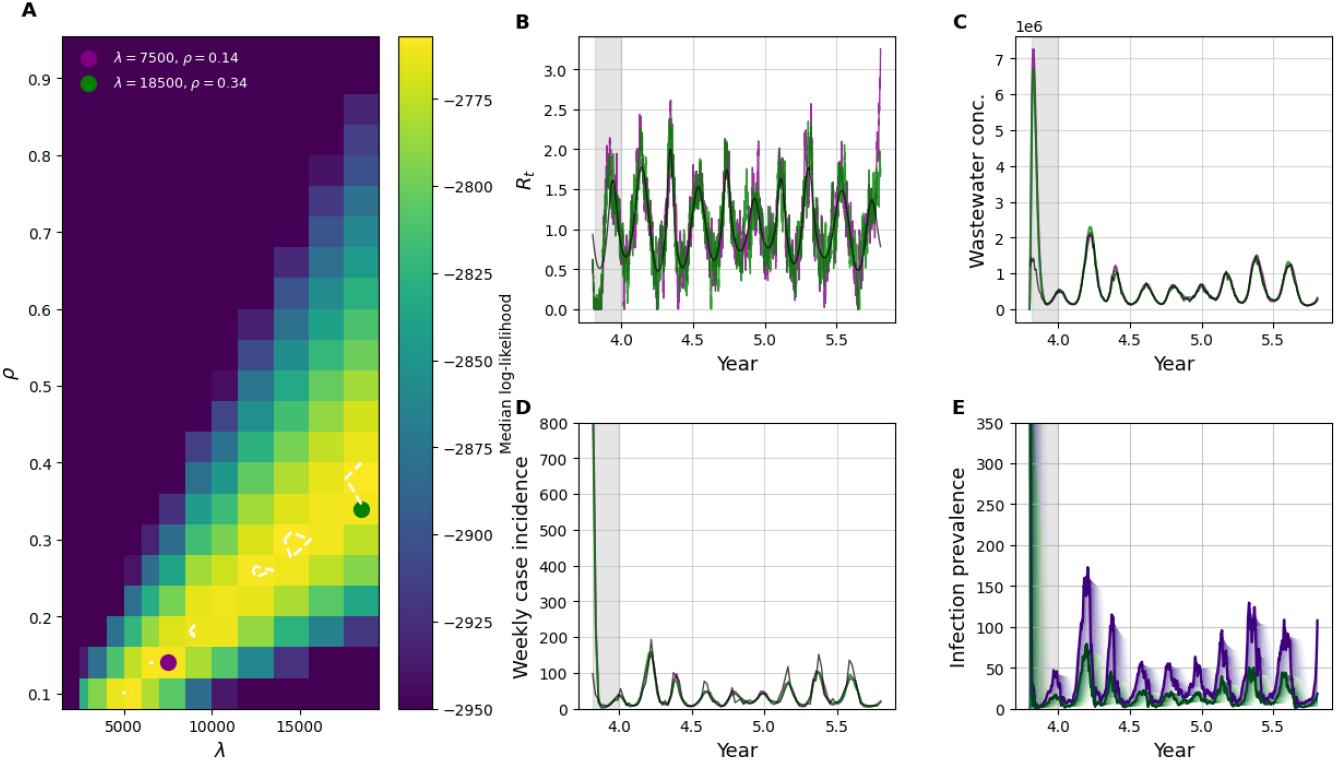
Estimation of the reporting rate *ρ* and the shedding load constant *λ* from the mock case and wastewater datasets. (A) Heatmap of log-likelihood values. Values below −2950 were set to −2950 for improved visualization. The green and purple dots mark two (*ρ, λ*) parameter combinations that lie along the log-likelihood ridge. We considered the first 70 days as a burn-in period and as such removed this period from the log-likelihood calculations. At each parameter combination, the log-likelihood value shown is the median value across 10 bootstrap particle filters. (B) Reconstructions of the time-varying effective reproduction number *R*_*t*_ under the two different (*ρ, λ*) parameter combinations shown with purple and green dots in panel A. True *R*_*t*_ values used to generate the mock data are shown alongside these reconstructed values. (C) Reconstructions of weekly case incidence under the two different (*ρ, λ*) parameter combinations, alongside the mock weekly case incidence data. (D) Reconstructions of wastewater concentration under the two different (*ρ, λ*) parameter combinations, alongside the mock wastewater concentration data. (E) Reconstructions of infection prevalence under the two different (*ρ, λ*) combinations shown with dots in panel A. The dynamics of *I*_0_ are shown in bold. The dynamics of higher-*i* infection classes are shown in lighter shades. In panels B-E, the 70-day burn-in period is shown with a gray band.

To further understand this result, we ran the bootstrap particle filter for two different (*ρ, λ*) combinations that lie along the observed log-likelihood ridge. Figure 6B shows their reconstructed *R*_*t*_ trajectories, Figure 6C shows their reconstructed wastewater concentrations, Figure 6D shows their reconstructed weekly case incidence, and Figure 6E shows their reconstructed prevalence. Both parameter combinations do a good job at reconstructing *R*_*t*_, observed wastewater concentrations, and case incidence data. Inferred infection prevalence is substantially higher when both *ρ* and *λ* are low than when these parameters have higher values (Figure 6E). These results indicate that identifiability of *ρ* and *λ* is not possible even when jointly fitting to case data and wastewater data. However, Figure 6A indicates that there are combinations of these parameters (high *ρ* and low *λ*; low *ρ* and high *λ*) that have much lower likelihoods and, as such, are unable to simultaneously reproduce observed patterns of case data and wastewater data. Together, these findings highlight that joint estimation of *R*_*t*_ from case incidence and wastewater concentrations impose strong constraints on the reporting rate and the shedding load scaling constant. Only combinations of parameters that fall within a high-likelihood region yield reconstructions that are consistent with both observed data sources.

### 3.2 Analysis of the Zurich Datasets

#### 3.2.1 *R*_*t*_ Estimation from the Zurich Datasets

Next, we applied our bootstrap particle filter to the SARS-CoV-2 case data and wastewater virus concentration data from Zurich over the time period September 3, 2020 through January 19, 2021. Figure 7A shows the log-likelihood surface across a range of Brown motion strengths (*σ*_*B*_ values) and across a range of reporting rates (*ρ* values) when the particle filter was applied to just the case data. This surface closely resembles the one from the mock case data, with *ρ* being unidentifiable and an intermediate value of *σ*_*B*_ = 0.32 yielding the highest likelihoods. In Figure 7B, we plot reconstructed *R*_*t*_ values for two parameter combinations: one with a low reporting rate (*ρ* = 0.2) and one with a high reporting rate (*ρ* = 0.8), both with *σ*_*B*_ = 0.32. Estimates of *R*_*t*_ reached up to 2.0 on October 7, 2020 and down to 0.5 on January 6, 2021. Both parameter combinations yielded similar estimates. Reconstructed daily incidence captured the observed case data under both parameterizations (Figure 7C).

**Figure 7.**
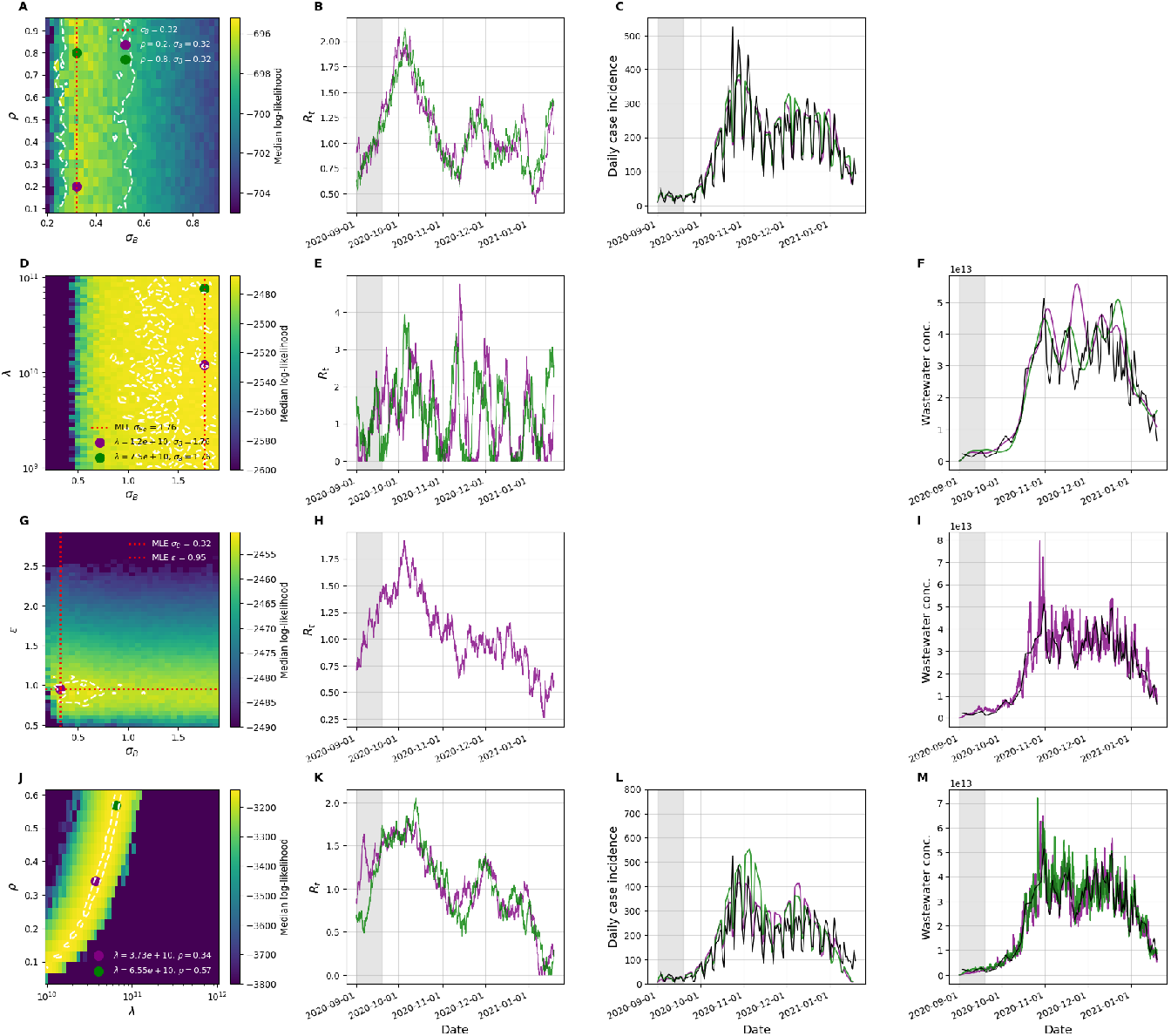
Parameter estimation and *R*_*t*_ inference for the Zurich datasets. (A)-(C) Estimation using Zurich case incidence data from September 3, 2020 to January 19, 2021. (D)-(F) Estimation using Zurich wastewater concentration data over the same time period without incorporating wastewater environmental noise. (G)-(I) Estimation using Zurich wastewater concentration data over the same time period, using a process model that incorporated environmental noise in viral outflow. (J)-(M) Estimation using both Zurich case incidence and wastewater concentration data over the same time period. Here, the process model incorporating environmental noise in viral outflow is used. (A, D, G) Heatmaps of log-likelihood values. Red dashed lines indicate maximum log-likelihood estimates. Green and purple dots mark two sets of parameter combinations whose reconstructed dynamics are shown in the panels further to the right. The first 18 days (gray bands in panels B, C, E, F, H, I, K, L, M) are removed from the log-likelihood calculations as a burn-in period. (B, E, H, K) Reconstructions of the time-varying effective reproduction number *R*_*t*_ under the two different parameter combinations shown with dots in panels A, D, G, and J, respectively. (C, L) Reconstructions of daily case incidence under the two different parameter combinations, alongside observed daily case numbers shown in black. (F, I, M) Reconstructions of wastewater virus concentrations under the two different parameter combinations, alongside observed wastewater virus concentrations shown in black.

Next, we applied our particle filter to just the Zurich wastewater virus concentration data. Figure 7D shows the log-likelihood surface across a range of shedding load constants (*λ* values) and *σ*_*B*_ values. Again, this log-likelihood surface resembles the one from the mock wastewater data, with *λ* being unidentifiable and a relatively high value of *σ*_*B*_ = 1.76 this time yielding the highest likelihoods. In Figure 7E, we plot reconstructed *R*_*t*_ values for two parameter combinations: one with a shedding load constant of *λ* = 1.2 *×* 10^10^ and one with a higher shedding load constant of *λ* = 7.5 *×* 10^10^, both with *σ*_*B*_ = 1.76. Estimates of *R*_*t*_ reached much higher values than they did for the case data (up to above 4.0) as well as much lower values than they did for the case data (hitting close to *R*_*t*_ = 0 multiple times over the time period studied). Given our analyses on the mock datasets, we know that the dramatic changes in *R*_*t*_ shown in Figure 7E are consequences of the high *σ*_*B*_ value used. Reconstructions of the wastewater virus concentrations under both the *λ* parameterizations were highly variable over time and differed between one another. Moreover, neither reconstruction was able to capture the high-frequency variation in the observed wastewater virus concentration data. These results indicate that the structure of the state space model that the particle filter is using may not be able to reproduce these wastewater data and therefore indicate to us that a modification of the process model may be needed.

We reasoned that the inability of the particle filter to capture the observed wastewater data stems from the high variability in wastewater measurements between temporally-adjacent samples. This variability can only be captured by the model (as is) through rapid and large changes in *R*_*t*_, which brings about a large amount of variation in the number of individuals infected across the different infected compartments. However, the case data do not point towards a large amount of variation in the number of infected individuals across compartments; beyond their day-of-week fluctuations, the case data vary relatively smoothly over time. We thus hypothesized that the rapid fluctuations in wastewater virus concentrations instead reflected the presence of environmental noise, specifically, variation in the rate at which virus is washed out or exits the sampled area. Variation in rainfall, for example, could impact this outflow rate (de Oliveira et al., 2021). To account for this, we introduced an environmental noise term by adding an additional stochastic component to the viral outflow process (see equation (15)).

Figure 7G shows the log-likelihood surface across a range of *ϵ* values and across a range of *σ*_*B*_ values, where the parameter *ϵ* quantifies the strength of environmental noise in viral outflow. The peak log-likelihood occurs at values of *σ*_*B*_ = 0.32 and *ϵ* = 0.95. This first indicates that the strength of Brownian motion for modeling *R*_*t*_ can be substantially lower when environmental noise in viral outflow from wastewater is incorporated into the process model. Indeed, the maximum likelihood estimate (MLE) of *σ*_*B*_ becomes similar to that of the MLE of *σ*_*B*_ when applying the particle filter to case data (Figure 7A). Figures 7H and 7I show reconstructed *R*_*t*_ and wastewater virus concentrations using the MLEs of *ϵ* and *σ*_*B*_. Compared to our results using a process model without environmental noise in viral outflow (Figure 7E), the inferred *R*_*t*_ trajectory (Figure 7H) appears less jagged and agrees more closely with that of the case data (Figure 7A). Moreover, the reconstructed wastewater virus concentrations successfully capture the smaller peaks and troughs present in the observed data (Figure 7I). The addition of environmental noise to the process model therefore considerably improved model fit.

Finally, we applied our particle filtering approach to both the case data and the wastewater data from Zurich. The state space model we applied incorporated environmental stochasticity in the viral outflow rate (*ϵ* set to 0.95) and set the strength of *R*_*t*_ Brownian motion to *σ*_*B*_ = 0.32, consistent with this estimate from Figures 7A and 7G. Figure 7J shows the log-likelihood surface across a range of *ρ* values and across a range of *λ* values. Here, as for the mock data (Figure 6A), there is a high-likelihood ridge spanning from low-*λ*/low-*ρ* parameter combinations to high-*λ*/high-*ρ* parameter combinations. Figures 7K, 7L, and 7M show reconstructed *R*_*t*_ trajectories, reconstructed case incidence, and reconstructed wastewater virus concentrations, respectively, under two parameter combinations that lie on this high-likelihood ridge (shown with dots in Figure 7J). These results again indicate that identifiability of *ρ* and *λ* is not possible even when jointly fitting to case data and wastewater data, although fitting to both datasets does impose strong constraints on possible parameter combinations that are consistent with observed data sources.

#### 3.2.2 Incorporating serological data to identify parameters and infection dynamics from the Zurich Datasets

Figures 4 and 5 indicate that the reporting rate *ρ* and the shedding load constant *λ* are not identifiable when using a bootstrap particle filter to fit to case data and wastewater virus concentration data, respectively. These figures further indicate that underlying infection dynamics are not identifiable (Figures 4D and 5D). Similarly, Figure 6 and Figures 7J-H show that the reporting rate and the shedding load constant are not identifiable when using a bootstrap particle filter to simultaneously fit to case data and wastewater virus concentration data, although only a subset of *ρ* and *λ* parameter combinations will yield high likelihoods (Figure 6A, 7J). Again, underlying infection dynamics are not identifiable, as a ridge of parameter combinations all have similar likelihoods despite different underlying infection dynamics. To summarize the different magnitudes of infection dynamics across these parameter combinations, we plot in Figure 8 the increases in seropositivity between September 15, 2020 and December 16, 2020. As expected (and consistent with Figures 4D and 5D), low-*λ*, low-*ρ* parameter combinations yielded the largest increases in seropositivity over this time span and high-*λ*, high-*ρ* parameter combinations yielded the smallest increases in seropositivity over this time span. In Figure 8, we further plot in red the empirical estimate of the change in seropositivity based on estimates from the two serological studies performed, which yielded an increase of 4.3% between these two dates. These serological studies, together with our bootstrap particle filter estimates, therefore indicate that the reporting rate is approximately *ρ* = 0.28 (28%) and that the shedding load constant is approximately *λ* = 3.1 × 10^10^ copies/mL. Serological studies can therefore resolve issues of parameter unidentifiability if conducted over an informative time period.

**Figure 8.**
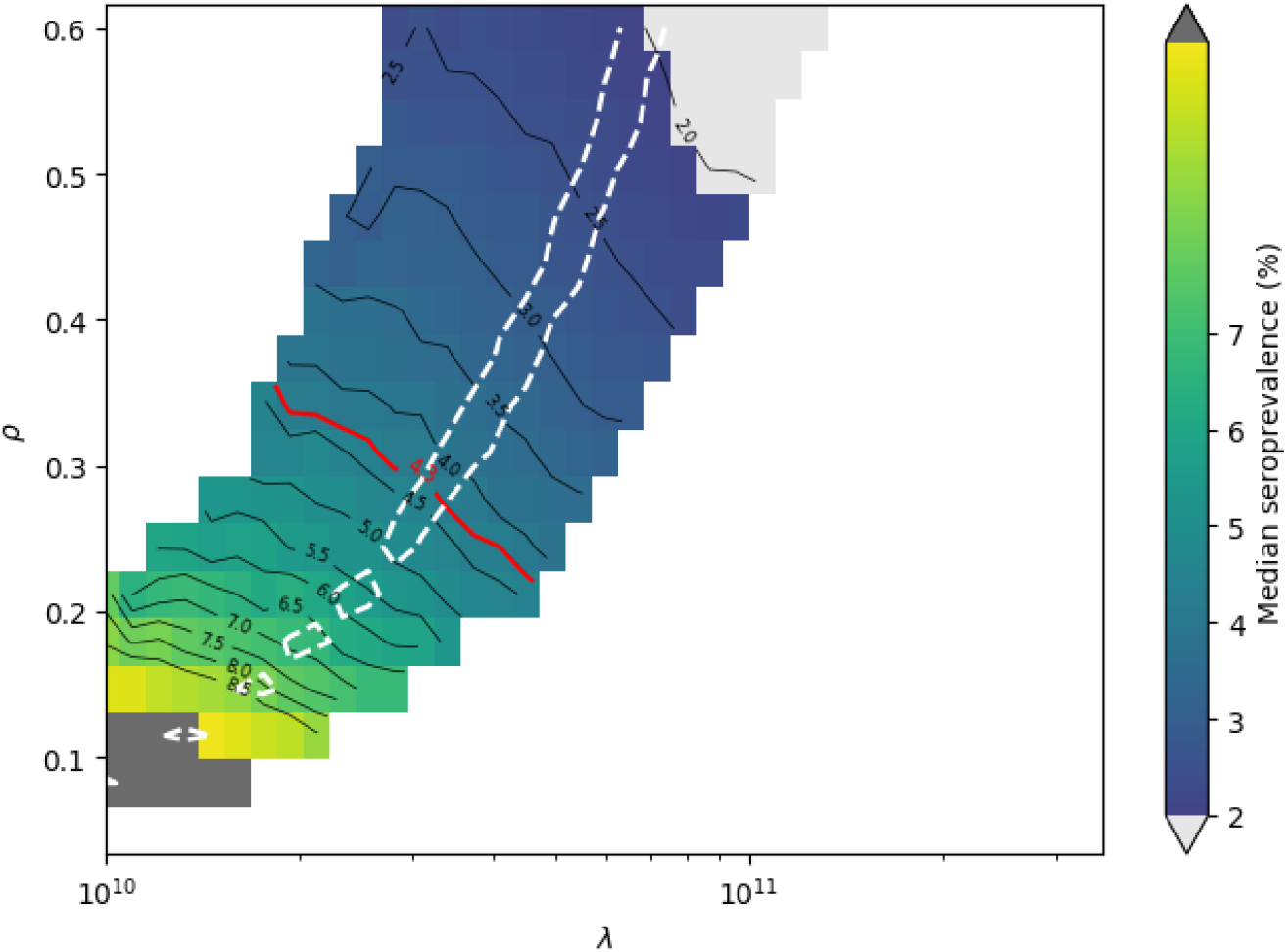
Heatmaps of seroprevalence changes between mid-September and mid-December across a range of *λ* and *ρ* parameter combinations. To generate the heatmap, 10 particle filter simulations were run at each parameter combination and the simulation with the median log-likelihood was chosen. Seroprevalence changes from these chosen simulations are shown in the heatmap. Estimates are based on Zurich case incidence and wastewater concentration data from September 3, 2020 to December 16, 2020. Seroprevalence changes are only shown for the subset of parameter combinations that yielded log-likelihood values less than or equal to –3350. The red dashed line is a contour line showing a seroprevalence change of 4.3%, as calculated from the available serological studies. The white lines show the 95% confidence region of the parameters.

#### 3.2.3 Forecasting the Zurich Data

Based on our analyses incorporating serological data (Figure 8), we next used our bootstrap particle filter, parameterized with *σ*_*B*_ = 0.32 and *ϵ* = 0.95, to forecast *R*_*t*_, daily case incidence, and wastewater virus concentrations. To do this, we first ran the bootstrap particle filter 10 times on the combination of Zurich case data and wastewater data, sampling 10 particles (1 from each run) at the last observation timepoint (January 19, 2021) to obtain *R*_*t*_, case incidence, and wastewater virus concentration reconstructions (Figure 9A,B,C, respectively). We then simulated these 10 sampled particles 10 days forward in time using the underyling process model including the Brownian motion of *R*_*t*_ and environmental noise implemented in the viral outflow rate. Figure 9A shows the *R*_*t*_ forecasts from these 10 particles. The particle forward simulations indicate that *R*_*t*_ values (estimated at *R*_*t*_ values between 0.5 and 1 on January 19, 2021) could reach as high as 1.5 or as low as 0.1 over the following 10 days. Despite this uncertainty in the trajectory of *R*_*t*_, forecasts of case incidence were bounded, with daily case incidence forecasted to lie between 50 and 150 daily cases within 10 days (Figure 9B). The observed daily case incidence data over this 10 day period fell squarely within our projections. Finally, the forecasts of wastewater virus concentrations were also bounded (Figure 9C), although we do not have wastewater virus concentration over this time period to assess the performance of these 10-day wastewater forecasts.

**Figure 9.**
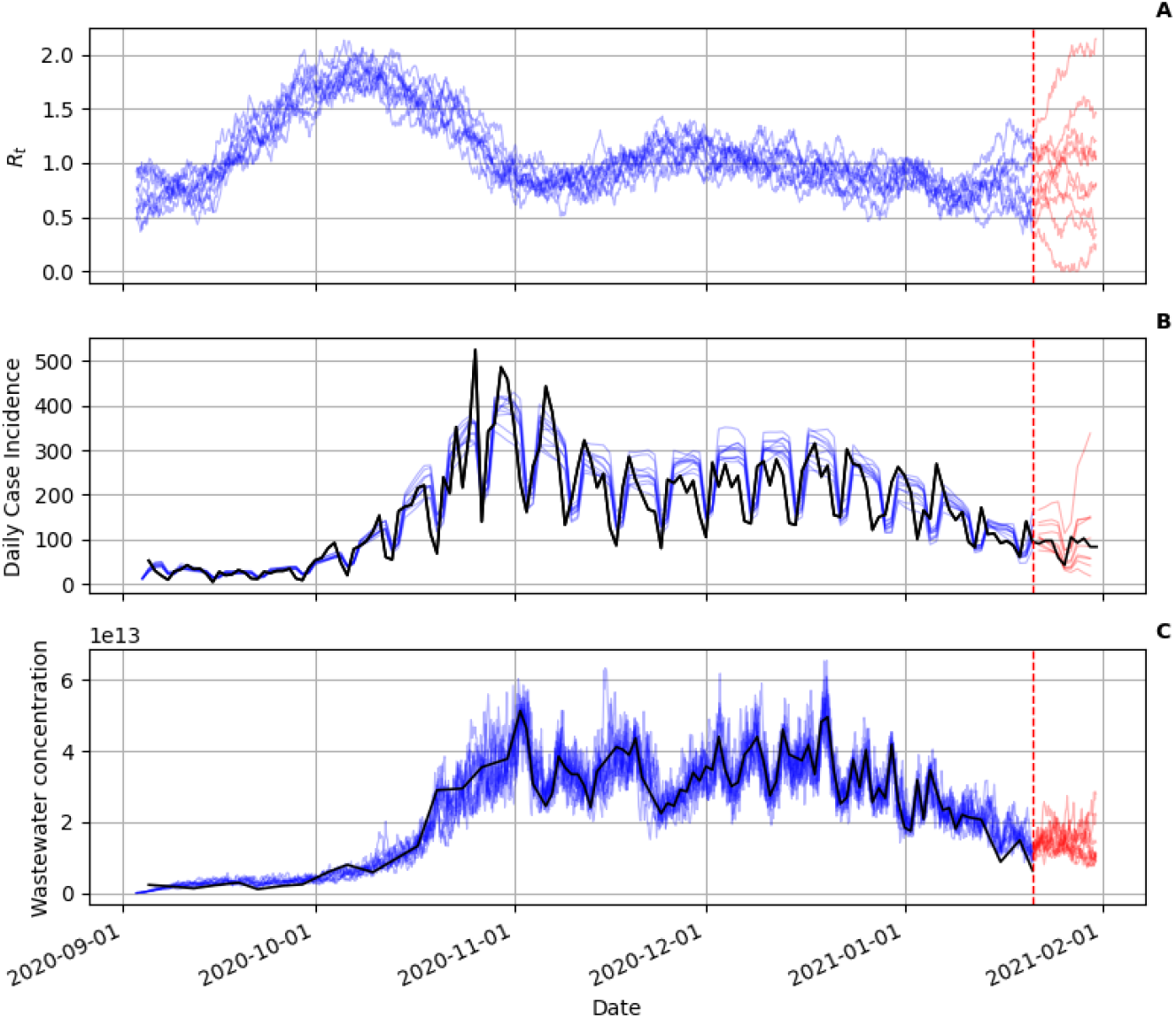
Forecasting *R*_*t*_, case incidence, and wastewater virus concentrations from the Zurich data. (A) Reconstructed *R*_*t*_ (blue) and forecasted *R*_*t*_ (red) from 10 sampled particles. Each particle was sampled from a different particle filter simulation. *R*_*t*_ was inferred using both case data and wastewater virus concentration data. (B) Reconstructed case incidence (blue) and forecasted case incidence (red) from the same 10 sampled particles as in panel A. Black lines show observed daily case incidence over the time period September 1, 2020-January 29, 2021. (C) Reconstructed wastewater virus concentrations (blue) and forecasted wastewater virus concentrations (red) from 10 sampled particles. In panels (A)-(C), the red dashed line shows the latest observation time point used by the particle filter.

## 4 Discussion

Wastewater data are starting to play an important role in understanding disease transmission and in guiding public health responses. In this study, we developed a simple epidemiological state space model and applied a bootstrap particle filter to this model to estimate the time-varying effective reproduction number *R*_*t*_ from case incidence data, wastewater virus concentration data, and then a combination of these two data streams. While particle filters have been extensively used to interface epidemiological models with case data (Ionides et al., 2006; Endo et al., 2019; Lemaitre et al., 2020), and even sequence data (Rasmussen et al., 2011, 2014a,b), their application to wastewater data has not been explored in great depth. Our state space model incorporated a process model that partitioned infected individuals into *n* compartments, which were accessed sequentially over time. Rates of infectiousness, case reporting, and viral shedding varied between compartments such that assumed infectiousness profiles, case reporting profiles, and shedding profiles reflected empirically-described distributions. To evaluate our approach, we generated a mock case dataset and a mock wastewater virus concentration dataset via forward simulation and applied our bootstrap particle filtering approach to these datasets. Our results indicated that the case reporting rate *ρ* was not identifiable when fitting only to case data but that nevertheless, one could use the particle filter to successfully reconstruct *R*_*t*_ and case data. Similarly, our results indicated that the shedding load constant *λ* was not identifiable when fitting only to wastewater data but that nevertheless, one could use the particle filter to successfully reconstruct *R*_*t*_ and wastewater virus concentration data. Finally, our results indicated that *ρ* and *λ* were not identifiable when fitting jointly to case data and wastewater data, although there were substantial constraints in the parameter combinations of *ρ* and *λ* that could successfully reconstruct *R*_*t*_ and the observed data streams.

We then applied our approach to COVID-19 case data and SARS-CoV-2 wastewater virus concentration data from Zurich, Switzerland. Inspection of the case data indicated that a day-of-week effect was present in the time series and we first modified our observation model to account for this effect. With this modification, we then attempted to estimate *R*_*t*_ and reconstruct case data and wastewater data. While we were able to estimate *R*_*t*_ and reconstruct case data, our application to wastewater data yielded highly variable *R*_*t*_ estimates that were inconsistent with *R*_*t*_ estimates from the case data. We speculated that this was a result of high day-to-day variability in wastewater virus concentrations that could only be captured in the current model formulation by dramatic fluctuations in *R*_*t*_. We therefore incorporated environmental stochasticity in the viral outflow rate into the process model to allow for an additional (non-*R*_*t*_) source of process noise. Once incorporated, we saw substantial improvements in our ability to estimate *R*_*t*_ from case data. This indicates to us that statistical approaches that integrate different sources of noise may more generally be needed to extract useful information from wastewater virus concentration data. Indeed, several studies have already highlighted different sources of noise in wastewater data (Lison et al., 2024; Keshaviah et al., 2023). Properly accounting for these various sources of noise in fitting models to data will, we think, continue to improve our understanding of disease dynamics from wastewater data.

Our analyses also showed that underlying infection dynamics were not identifiable from case data or wastewater data or from their combination alone. However, we showed that by considering serological data that we were able to then identify reporting rates and shedding load constants, and therewith also resolve underlying infection dynamics. Our approach here did not rely on explicitly incorporating serological data into our inference approach; we simply added a variable (*S*) to allow us to keep track of changes in seropositivity and used simulated changes in this variable between the two serological testing dates to identify the best set of plausible parameter combinations that yielded high likelihoods when interfaced with case data and wastewater virus concentration data. Incorporating serological data as an additional data source during the inference process is possible and future work should explore this possibility, especially with longer time series and in the case of multiple serological data points. In the case of longer time series, it will be necessary to include the process of seroreversion (by setting the parameter *γ*_*S*_ to a positive value based on empirical estimates (e.g., those from Shioda et al. (2021)). Note that although we showed that serological data are useful in allowing us to identify infection dynamics when both case data and wastewater data were available, our analyses shown in Figures 4 and 5 also indicate that serological data would allow us to identify infection dynamics if *only* case data or wastewater data were available as data streams. Finally, we showed that the bootstrap particle filter could also be used for short-term forecasting of *R*_*t*_, case numbers, and wastewater virus concentrations. This work therefore adds to existing studies that have aimed to forecast disease dynamics using wastewater data (Schenk et al., 2024; Manuel et al., 2024; Cañas Cañas et al., 2025; Radermacher et al., 2025).

Our work also contributes to the growing body of approaches that leverage wastewater data for infectious disease surveillance with the aim of reconstructing *R*_*t*_. Given the increase in the number of approaches, recent work has also started, to evaluate the relative performance of different approaches (Hill et al., 2025). One set of approaches relies on discrete-time convolutions to first infer underlying incidence time series from wastewater data (assuming a given shedding load distribution) and then to apply the Cori et al. (2013) estimation approach to these inferred incidence time series to reconstruct *R*_*t*_ (Huisman et al., 2022; Amman et al., 2022; Wannigama et al., 2023). However, these approaches do not handle missing data well nor can they easily incorporate additional data streams such as case data. In contrast, particle filtering methods, such as the one proposed here, do not suffer from these issues. Notably, another approach combines reported cases with wastewater data in a semi-mechanistic model that uses particle Markov Chain Monte Carlo (pMCMC) to estimate *R*_*t*_ and a time-varying case ascertainment rate (Watson et al., 2024). Our model, similar to the pMCMC method, is designed to integrate multiple datasets, including case incidence and wastewater viral concentration data, without the need for imputing missing data. In comparison to the Watson et al. (2024) approach, our approach instead uses a Markovian process model, which allows for a flexible incorporation of additional sources of environmental noise (such as temporal variation in viral outflow rates).

Despite the advantages of our approach, it is not without limitations. For example, for SARS-CoV-2, studies have shown that as little as 2% of cases can be responsible for 20% of infections, with such heterogeneity being age-specific (Lau et al., 2020). Our process model does not include demographic stochasticity and thereby ignores this transmission heterogeneity. Moreover, substantial heterogeneity exists in the extent of virus shedding between individuals (Ke et al., 2022) and our model also ignores this heterogeneity, which may be important to include when considering small sewer catchment areas. Our model also does not consider the impact of variants of concern on transmission and virus shedding. In particular, using a constant shedding load scaling constant *λ* and a fixed shedding load distribution overlooks described variant-specific differences in shedding rates (Prasek et al., 2023). Changes in immunity or differences in the relative infectiousness of variants are also not taken into account by the model. Furthermore, the model’s predictions rely on the assumption that *R*_*t*_ changes according to a Brownian motion model. As a result, the model will not respond immediately to rapid changes in *R*_*t*_ driven by interventions such as social distancing, mask mandates, or school closures. Finally, without explicit incorporation of seasonal forcing, the model has limited ability to capture regular, longer-term trends, such as seasonal dynamics over the course of a year, particularly for pathogens like influenza.

Our model structure also adopted several assumptions and incorporated several decisions that could be reconsidered and modified in other applications. One such decision was that *R*_*t*_ changed according to a Brownian motion model, rather than *log*(*R*_*t*_) changing according to a Brownian motion model. In our analyses of mock datasets, we found that inference assuming that *R*_*t*_ changes according to a Brownian motion model performed better than inference assuming that *log*(*R*_*t*_) changing according to a Brownian motion model. However, this choice put some biological constraints on the Brownian motion model. Specifically, when a particle’s *R*_*t*_ value became negative, we decided to reset *R*_*t*_ to be zero. An alternative approach could be to use a reflective boundary, setting *R*_*t*_(*t*+1) to |*R*(*t*) +*dR*_*t*_|. Our equation (3) for case data also overcounts the number of cases when *ρ* is large. This is because a single individual could be double-counted as a case as they transition through the various *I* infection classes. One way to work around this issue would be to expand the model to include two separate classes of infected individuals: those who have not been detected as cases and those who have already been detected as cases. At small values of *ρ*, this extended model with a parallel chain would reduce to the single-chain model we used here. While this extension would make our inference of *ρ* exact, it would double the number of infected compartments and therefore be more computationally intensive. As our primary interest here was in reconstructing *R*_*t*_, we therefore decided to keep with the simpler model. The structures and parameterizations of our observation models also necessarily adopted some assumptions that could be modified. For example, we used a negative binomial distribution parameterized with low overdispersion (high *k*_*c*_) as our observation model for cases (equation (11)). We could have instead used a Poisson distribution or alternative distribution or increased the amount of overdispersion by choosing a lower value of *k*_*c*_. For the wastewater observation model, we used a Gamma distribution (equation (12)). We could have alternatively used a Student’s *t*-distribution as was done in Goldstein et al. (2023). In application to the Zurich dataset, this may have allowed us to keep the major January 10, 2021 outlier in the dataset, which was supported by both N1 and N2 measurements but exceeded temporally adjacent timepoints by over 160%. While we used a Gamma distribution in our analysis, we did attempt to show the flexibility of using this distribution by assuming that the coefficient of variation was a constant value when fitting to the mock data and by instead assuming that the variance of this distribution was a constant value when fitting to the Zurich wastewater data.

Despite the simplifying assumptions detailed above and the necessary choice of adopting specific process model and observation model structures, we here showed that we could statistically interface our state-space model with case incidence data and/or wastewater virus concentration data using a bootstrap particle filter to successfully reproduce time-varying effective reproduction numbers from a mock data set. Our approach is both fast (with a particle filter run taking less than 3 seconds to execute) and versatile in that infection profiles, case detection profiles, and shedding profiles can be easily modified for another pathogen. Given these benefits, we hope that our approach will prove useful in contributing to effective, data-driven public health decision-making.

## Code and data availability

All code and mock datasets that are needed to reproduce the results in this paper can be found at: https://github.com/koellelab/Rt_wastewater_inference_particle_filter.

## Acknowledgments

We thank members of the Koelle lab and InsightNet for feedback on this work.

